# Beyond the Red Complex: *De Novo* Marker Discovery Uncovers Novel Periodontitis-Associated Taxa and Enables Non-Invasive Machine Learning Diagnosis

**DOI:** 10.64898/2026.07.31.26359396

**Authors:** Mohamad Koohi-Moghadam, Wai Keung Leung

## Abstract

**Background:** Periodontitis affects over 1 billion people worldwide, yet diagnosis relies on clinical measures that capture tissue destruction rather than underlying microbial dysbiosis. Most microbial-biomarker studies use 16S rRNA sequencing or reference-database mapping, systematically under-detecting uncultivated or divergent taxa.

**Methods:** We assembled 341 supra- and subgingival shotgun metagenomes (218 periodontitis, 123 health) across nine countries/regions. Using MetaMarker, a de novo, reference-free pipeline, we identified conserved genomic markers directly from reads in a 305-sample discovery pool without database mapping. Markers were taxonomically annotated against the Human Oral Microbiome Database, functionally annotated with Prodigal/eggNOG-mapper, and used for eight machine-learning classifiers, externally validated on three independent held-out cohorts (36 samples).

**Results:** We recovered 2,142 significant markers (1,999 periodontitis-enriched, 143 health-enriched; q<0.05), recapitulating the canonical red/orange-complex dysbiotic shift. Beyond established pathogens, 128 periodontitis markers (6.4%) were novel, including an uncultivated Paludibacteraceae genus and divergent Fretibacterium fastidiosum strains. Case markers encoded a coherent virulence programme spanning proteolysis, haem/iron acquisition, and Type IX secretion. On external validation, boosted-tree classifiers generalized best (XGBoost and gradient boosting, AUC=0.96), and a compact SHAP-ranked 20-marker panel spanning five taxa reproduced full-set directionality.

**Conclusions:** Reference-free metagenomic marker discovery recovers known periodontal pathobiology while revealing unrecognized candidate biomarkers and supports an accurate, externally validated, non-invasive classifier with translational potential as a compact diagnostic panel.

## 1. Introduction

Periodontitis is a prevalent oral disease characterized by an imbalance in the host’s immunoinflammatory responses to dysbiotic oral biofilms, ultimately leading to the destruction of periodontal tissues [1]. The 2021 Global Burden of Disease Study reported that over 1 billion people were affected by severe periodontitis in 2021, and by 2050 this number is projected to rise to more than 1.5 billion, representing a 44.32% increase [2]. Diagnosis has traditionally relied on clinical measurements such as probing depth, attachment loss, and bleeding on probing [3]; while indispensable, these measures capture the consequences of tissue destruction rather than the underlying dysbiotic process and offer limited resolution for early or subclinical disease.

Periodontitis is fundamentally a polymicrobial disease: subgingival dysbiosis is dominated by the so-called “red complex” (*Porphyromonas gingivalis*, *Tannerella forsythia*, *Treponema denticola*) and an expanding set of “orange complex” and emerging pathobionts, acting in concert rather than as single etiological agents [4–6]. This has motivated growing interest in molecular biomarkers as adjuncts or alternatives to clinical probing, with proteins such as IL-1, MMP-8, and ICTP measured in saliva as indicators of periodontal host response [7, 8]. In parallel, next-generation sequencing (NGS) has enabled identification of dysbiosis-associated microbial signatures, with specific taxa—*P. gingivalis*, *T. denticola*, *T. forsythia*, and more recently *Prevotella intermedia*, *Campylobacter rectus*, and *Fusobacterium nucleatum*— proposed as microbial biomarkers for early detection [9–11]. However, most prior microbial-biomarker studies have relied on 16S rRNA amplicon sequencing or reference-database-mapped shotgun profiling, both of which are fundamentally constrained by the composition of existing reference collections: uncultivated, divergent, or newly described taxa are systematically under-represented or missed entirely, and strain-level genomic information is collapsed or discarded [12, 13].

Whole-metagenome shotgun sequencing offers substantially higher taxonomic and functional resolution than 16S profiling, but conventional analysis pipelines still typically map reads to a reference database (e.g., to build an OTU or species abundance table), thereby inheriting the same database bias and losing sequences that do not match any reference genome. An alternative is de novo, reference-free marker discovery, in which candidate biomarkers are defined directly from the read data as conserved genomic fragments that differentiate disease groups, independent of any prior reference database. This concept was introduced by MetaMarker [14], a de novo approach that does not require mapping raw reads to a reference database, with biomarkers found by MetaMarker being conserved sequence fragments that are shared by a subgroup of bacteria, and previously validated for colorectal cancer biomarker discovery and classification.

In this study, we assembled a large, multi-cohort collection of supra- and subgingival shotgun metagenomes—341 samples drawn from seven independent studies across the United States, Italy, Hong Kong, China, Japan, Belgium, Chile, Peru, and Spain—to identify novel, reference-free microbial biomarkers for periodontitis. Using MetaMarker, a de novo marker-discovery pipeline, we identified conserved genomic domains as candidate biomarkers directly from the sequencing reads, annotated their taxonomic origin and encoded function, and developed a machine-learning classifier trained on marker abundance and evaluated on three fully independent, held-out external cohorts. Our specific aims were to: (i) determine whether de novo markers recapitulate known periodontal microbiology; (ii) identify candidate biomarkers from taxa not previously associated with periodontitis; (iii) characterize the functional/virulence programme encoded by disease-associated markers; and (iv) build and externally validate a transferable, interpretable classifier—including a compact, clinically actionable marker panel—for non-invasive periodontitis diagnosis.

## 2. Materials and Methods

### 2.1 Study cohorts and data

We assembled a multi-cohort collection of 341 supra- and subgingival shotgun metagenomic samples (218 periodontitis, 123 health/control) from 11 publicly available BioProjects spanning nine countries/regions. Seven studies (PRJNA552294, PRJNA508385, PRJNA255922, PRJNA78025, PRJNA48479, PRJNA1238390, PRJDB11203, and PRJNA1183294) were pooled for marker discovery, abundance quantification, and classifier training. Three independent studies (PRJNA547717, Italy; PRJNA932553, Hong Kong; PRJCA003936, China; 36 samples total: 17 periodontitis, 19 health) were withheld in their entirety as an external test set and contributed no data to marker discovery, abundance ranking, or classifier training (**Table 1, Supplementary Table 1**).

**Table 1.**
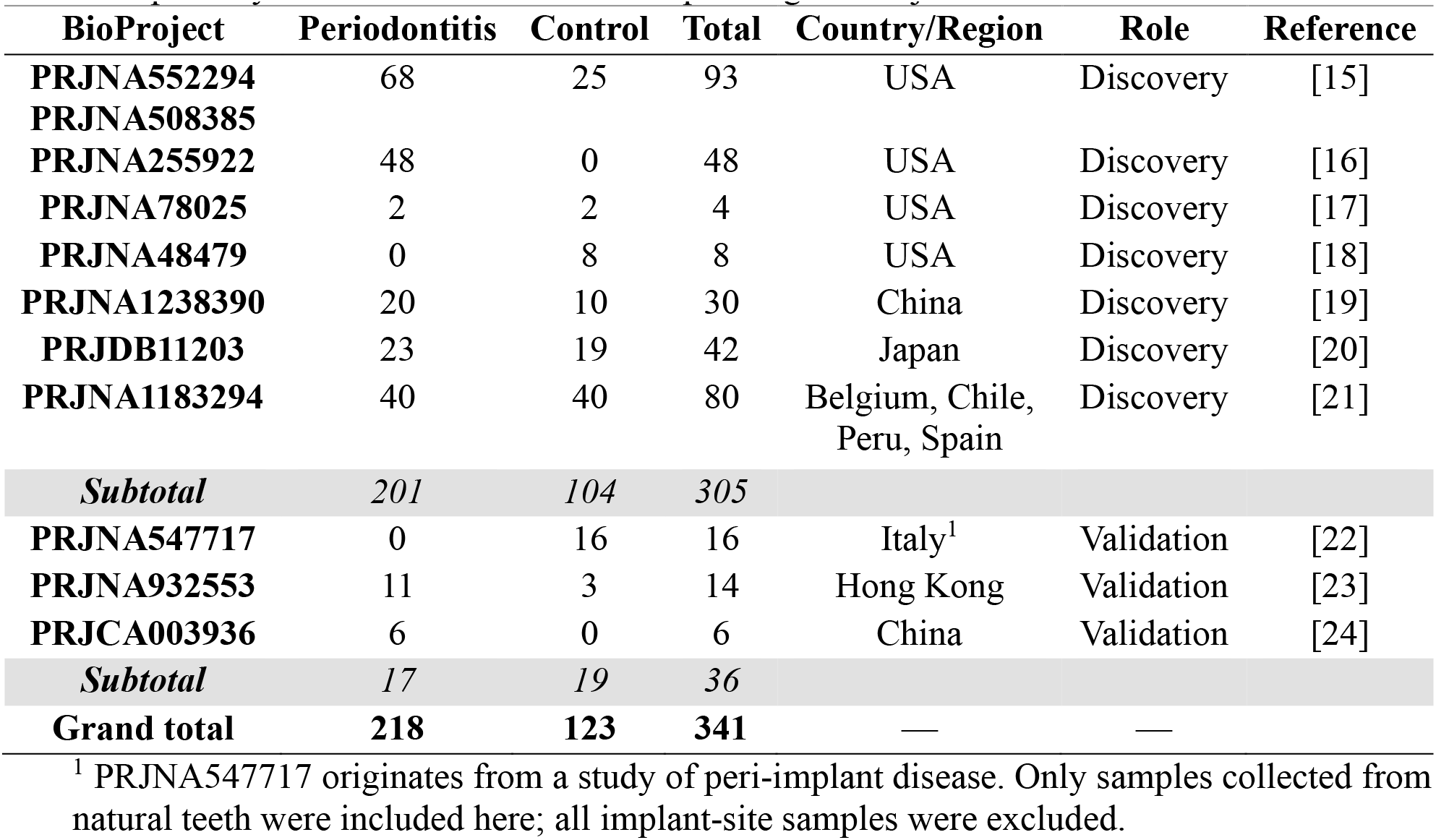
Study cohorts used in this work. Sample counts reflect only those samples that were publicly available within each corresponding BioProject at the time of data collection.

| **BioProject** | **Periodontitis** | **Control** | **Total** | **Country/Region** | **Role** | **Reference** |
| --- | --- | --- | --- | --- | --- | --- |
| **PRJNA552294** | 68 | 25 | 93 | USA | Discovery | [15] |
| **PRJNA508385** |  |  |  |  |  |  |
| **PRJNA255922** | 48 | 0 | 48 | USA | Discovery | [16] |
| **PRJNA78025** | 2 | 2 | 4 | USA | Discovery | [17] |
| **PRJNA48479** | 0 | 8 | 8 | USA | Discovery | [18] |
| **PRJNA1238390** | 20 | 10 | 30 | China | Discovery | [19] |
| **PRJDB11203** | 23 | 19 | 42 | Japan | Discovery | [20] |
| **PRJNA1183294** | 40 | 40 | 80 | Belgium, Chile,<br>Peru, Spain | Discovery | [21] |
| ***Subtotal*** | ***201*** | ***104*** | ***305*** |  |  |  |
| **PRJNA547717** | 0 | 16 | 16 | Italy<sup>1</sup> | Validation | [22] |
| **PRJNA932553** | 11 | 3 | 14 | Hong Kong | Validation | [23] |
| **PRJCA003936** | 6 | 0 | 6 | China | Validation | [24] |
| ***Subtotal*** | ***17*** | ***19*** | ***36*** |  |  |  |
| **Grand total** | **218** | **123** | **341** | — | — |  |

<sup>1</sup> PRJNA547717 originates from a study of peri-implant disease. Only samples collected from natural teeth were included here; all implant-site samples were excluded.

### 2.2 Read preprocessing and host removal

Whole-metagenome shotgun reads were processed directly from their native format; both FASTA and FASTQ inputs (plain or gzip-compressed) were accepted without prior conversion. Prior to marker discovery, all FASTQ reads underwent quality control with fastp v0.23.4: low-quality bases were trimmed from read ends (Phred quality threshold Q20), adapter sequences were removed using fastp’s built-in overlap-based adapter detection, and reads shorter than 50 bp after trimming were discarded. Per-sample quality-control summary statistics (raw/filtered read counts, mean quality, adapter content) were retained for downstream QC review. Marker discovery was subsequently performed on the quality-filtered reads that were retained after this step. Because sub- and supragingival metagenomes contain host DNA, human sequence was removed before marker de-replication. Assembled per-sample contigs were aligned to the human reference genome (GRCh38/hg38) with minimap2 v2.31 (asm10 preset), and any contig for which ≥50% of its length was covered by host alignments at ≥90% nucleotide identity was discarded as host-derived. The minimap2 host index was built once and reused across all samples.

### 2.3 De novo marker discovery

We used MetaMarker v2.0 [14] to identify conserved sequence markers directly from raw reads without mapping to a reference database, using canonical k-mer profiling, de Bruijn graph-based marker assembly, and clustering-based de-replication. The abundance of every retained marker was quantified in each of the 305 samples in our discovery set by mapping reads to the marker set with Bowtie2 v2.5.5. Per-marker counts were normalized to hits per million reads (HPM). Markers were ranked for differential abundance using Mann–Whitney U test on per-sample HPM (greater for periodontitis-enriched “Case” markers; lower for health-enriched “Control” markers). Benjamini–Hochberg (BH) false-discovery-rate q-values are reported for all markers. This yielded 1,999 Case and 143 Control markers (2,142 total). Full algorithmic details are provided in the original publication [14].

### 2.4 Taxonomic annotation

Each marker was assigned a source organism by nucleotide alignment with BLAST+ blastn v2.17.0 against a local database built from the Human Oral Microbiome Database (HOMD) [25] reference genome collection. Alignments used an E-value threshold of 1×10⁻¹⁰ and up to five target sequences per query; the highest-scoring hit (by bitscore) determined the marker’s species/strain assignment, along with percent identity and query coverage.

### 2.5 Strain-conservation analysis

To assess whether markers represent strain-conserved genomic regions rather than strain-specific artifacts, the top 20 markers by classifier model importance were aligned with NCBI blastn (megablast) against the core_nt database. For each marker, per-position nucleotide identity was computed across qualifying subject strains (query coverage > 90%, ten best hits shown; or a relaxed >50% coverage/top-3 threshold for markers too long for any single genome to span), and windowed identity profiles (50-bp windows) were plotted along the marker length.

### 2.6 Functional annotation and enrichment

Protein-coding genes were predicted on marker contigs with Prodigal v2.6.3 [26] (metagenomic mode). Predicted proteins were functionally annotated with eggNOG-mapper v2.1.13 [27] against the eggNOG 5.0 orthology database, using DIAMOND v2.2.2 [28] (blastp; --sensitive --iterate, E-value ≤1×10⁻³); each gene was assigned KEGG orthologs/pathways/modules, GO terms, COG functional categories, EC numbers, and Pfam/CAZy domains. Functional over-representation of Case-versus Control-marker genes was assessed for each KEGG pathway, COG category, and GO term using Fisher’s exact test with BH-adjusted FDR. Case-marker genes were additionally screened for periodontitis-relevant virulence functions (proteolysis, heme/iron acquisition, Type IX secretion, LPS/lipid-A biosynthesis, adhesion/fimbriae, motility, toxins, antibiotic resistance) by keyword matching over gene descriptions, preferred names, Pfam domains, and EC numbers.

### 2.7 Machine-learning models

Marker abundances were used to train binary classifiers (periodontitis vs. health). Features were the 2,142-marker HPM values, transformed as log₁₀(HPM+1) and standardized with a StandardScaler fit on training data only (embedded in a scikit-learn Pipeline). Eight model families were compared: L2-regularized logistic regression [29], RBF-kernel SVM [30], k-nearest neighbours (k=7) [31], random forest [32], extra-trees [33], gradient boosting [33], XGBoost (eXtreme Gradient Boosting) [34], and LightGBM (Light Gradient Boosting Machine) [35] using scikit-learn v1.7.2.

Studies were partitioned so that the three fully independent external cohorts (PRJNA547717, PRJCA003936, PRJNA932553; 36 samples: 17 periodontitis, 19 health; Table 1) were held out entirely as an unseen test set, while the remaining 305 samples across seven studies formed the training pool. Generalization within the training pool was estimated by stratified 5-fold cross-validation; final performance was reported on the held-out studies. Models were evaluated by AUC, accuracy, F1, precision, recall, and average precision. Model interpretability for the three best models was assessed by SHapley Additive exPlanations (SHAP).

### 2.8 Statistics, software, and data availability

Statistical tests were two-sided unless a directional hypothesis is stated; multiple testing was controlled by the Benjamini–Hochberg procedure (q < 0.05). A fixed random seed (random_state = 42) was used for all stochastic steps to ensure reproducibility.

## 3. Results

### 3.1 De novo markers recapitulate known periodontal microbiology

We recovered 2,142 non-redundant sequence markers that distinguished periodontitis from health without recourse to any reference database: 1,999 markers enriched in periodontitis (“Case” markers) and 143 enriched in health (“Control” markers). Across all 305 samples, all 2,142 markers were significant at a Benjamini–Hochberg FDR of q < 0.05 (Mann–Whitney U) (**Supplementary Tables 2 and 3).**

Hierarchical clustering of the 80 most discriminative markers (top 40 Case + top 40 Control by q-value) across 305 samples produced a clear block (“checkerboard”) structure in which periodontitis and health samples separated into distinct clusters along the top dendrogram, with Case markers—dominated by *Treponema denticola*, *Porphyromonas gingivalis* and *Tannerella forsythia*—elevated in periodontitis samples, and Control markers—dominated by *Streptococcus mitis*, *Streptococcus oralis*, and *Rothia dentocariosa*/*aeria*—elevated in health samples (**Figure 1**). This block structure was consistent across the multiple cohorts represented in the sample set, indicating that the discriminative signal is not driven by a single study.

**Figure 1.**
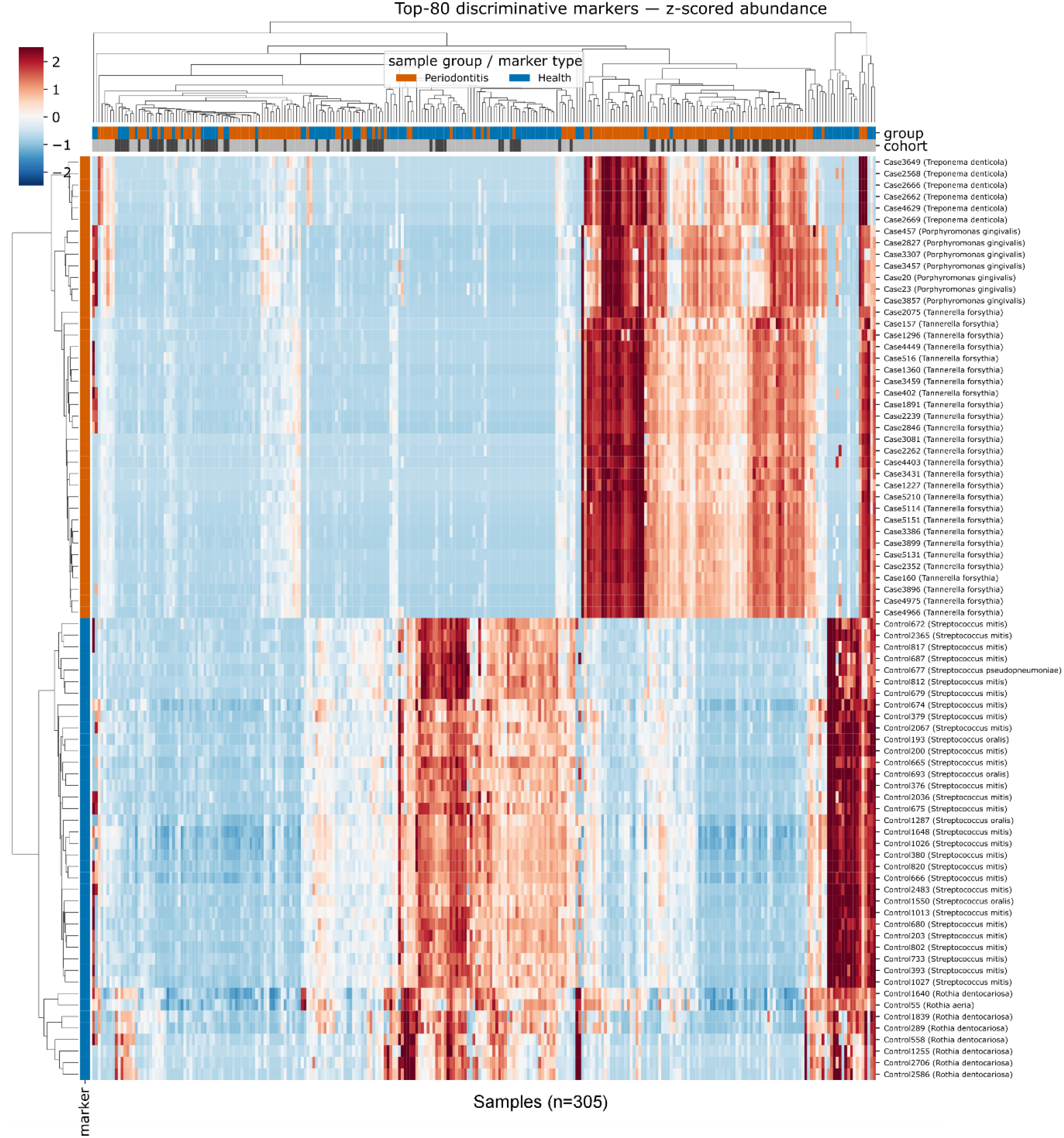
Hierarchical clustering of the top 80 discriminative markers (40 Case + 40 Control) across 305 samples. Rows are z-scored marker abundance, annotated by marker type (Case/Control) and source taxon; columns are samples, annotated by clinical group (periodontitis/health) and cohort. The checkerboard block structure shows coordinated elevation of red-complex/pathobiont markers in periodontitis and commensal markers in health.

At the level of individual markers, the top-ranked periodontitis-enriched and health-enriched markers each cleanly separated the two groups. The strongest periodontitis-enriched markers derived from *Treponema denticola* (log2FC = +3.2, q = 9×10⁻²¹), *Tannerella forsythia* (log2FC = +2.9, q = 2×10⁻²⁰), and *Porphyromonas gingivalis* (log2FC = +2.9, q = 9×10⁻²¹), all showing near-absence in health and marked, wide-ranging elevation in periodontitis. Conversely, the strongest health-enriched markers derived from *Rothia dentocariosa* (log2FC = −1.9, q = 2×10⁻¹⁶), *S. mitis* (log2FC = −2.7, q = 6×10⁻¹⁶) and *S. oralis* (log2FC = −2.4, q = 9×10⁻¹⁵), which were abundant in health but depleted in periodontitis (**Figure 2A**).

**Figure 2.**
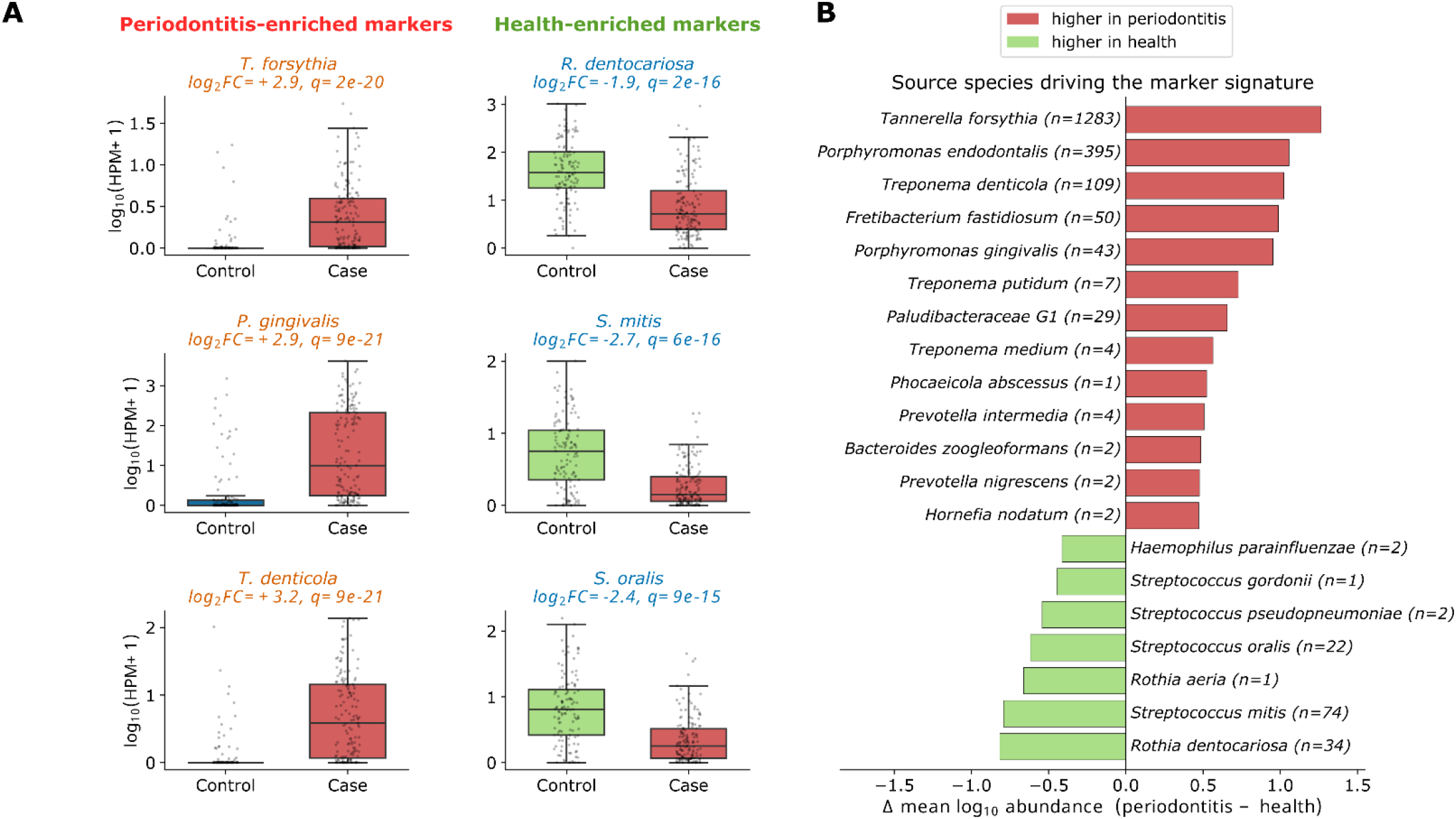
Individual markers and source organisms driving the periodontitis signature. (A) Boxplots of log₁₀(HPM+1) abundance in Control (green) vs. Case (red) samples for representative periodontitis-enriched markers (*Tannerella forsythia*, *Porphyromonas gingivalis*, *Treponema denticola*) and health-enriched markers (*Rothia dentocariosa*, *Streptococcus mitis*, *Streptococcus oralis*), annotated with log2 fold-change and BH q-value. (B) Marker abundance aggregated per source organism, showing mean difference in log₁₀ abundance (periodontitis − health) for the leading taxa driving the periodontitis (red) and health (green) signatures.

Aggregating marker abundance to the level of source organism confirmed that this pattern held genome-wide rather than being limited to a handful of top markers. Species elevated in periodontitis were the established red/orange-complex pathogens and emerging pathobionts— *Tannerella forsythia* (1,283 markers), *Porphyromonas endodontalis* (395), *Treponema denticola* (109), *Fretibacterium fastidiosum* (50), *Porphyromonas gingivalis* (43), *Treponema putidum* (7), the uncultivated *Paludibacteraceae* G1 (29), *Treponema medium* (4), *Prevotella intermedia* (4), and additional low-prevalence taxa including *Phocaeicola abscessus* (1), *Bacteroides zoogleoformans* (2), *Prevotella nigrescens* (2) and *Hornefia nodatum* (2)— whereas species elevated in health were oral commensals, principally *Rothia dentocariosa* (34 markers), *S. mitis* (74), *S. oralis* (22), *Rothia aeria* (1), *Streptococcus pseudopneumoniae* (2), *Streptococcus gordonii* (1) and *Haemophilus parainfluenzae* (2) (**Figure 2B**). The *de novo* markers therefore reconstruct the canonical dysbiotic shift of periodontitis from first principles, independent of any prior taxonomic reference.

### 3.2 Functional characterization of the discriminatory markers

Predicted proteins were called on the discriminatory marker contigs with Prodigal and functionally annotated with eggNOG-mapper. Gene calling yielded 5,309 open reading frames from the 1,999 Case markers and 355 from the 143 Control markers, of which 5,011 (94.4%) and 344 (96.9%), respectively, received a functional annotation (**Supplementary Tables 4 and 5).**

The two marker sets were functionally and taxonomically distinct. Control markers, derived almost exclusively from early-colonizing commensals of healthy plaque, were significantly enriched (Fisher’s exact, BH q < 0.05) for housekeeping and saccharolytic functions: ribosomal proteins/translation (COG J, q = 2.5×10⁻⁷), carbohydrate transport (COG G, q = 3.3×10⁻³), and the KEGG pathways ribosome, ABC transporters, PTS, and starch/sucrose and amino-sugar metabolism (**Table 2**). Case markers, originating overwhelmingly from *T. forsythia*, *P. endodontalis*, *T. denticola*, *P. gingivalis*, *F. fastidiosum*, and *Filifactor alocis*, were significantly enriched only for cell-envelope biogenesis (COG M, q = 7.5×10⁻³) and genes of unknown function (COG S, q = 5.7×10⁻³) at the coarse COG level — reflecting the sparser characterization of these anaerobes relative to well-studied commensals — with no individual KEGG orthologue reaching significance in the direct comparison. We therefore characterized the periodontitis markers by their intrinsic gene content rather than by differential enrichment alone.

**Table 2.** Housekeeping and saccharolytic functions encoded by the health-associated (control) markers (complete list in Supplementary Table 6).

| Functional category | Genes | Markers | Taxa | Representative genes | Principal contributing taxa |
| --- | --- | --- | --- | --- | --- |
| Translation & ribosome biogenesis | 52 | 31 | 5 | *rplN, rpsB, rpsL, tsf, gidA/mnmA* | *S. mitis* (29),<br>*S. oralis* (11),<br>*R. dentocariosa* (8) |
| ABC transporters & PTS system | 37 | 23 | 3 | *ptsG/ptsH/ptsI, manM/manN/manL/manZ, msmK, pstB1/pstB2 (phosphate ABC)* | *S. mitis* (27),<br>*S. oralis* (8),<br>*R. dentocariosa* (2) |
| Carbohydrate & sugar metabolism (saccharolytic enzymes) | 21 | 13 | 3 | *pulA2, treC/malQ, fba, pgi, agaS* | *S. mitis* (14),<br>*R. dentocariosa* (4),<br>*S. oralis* (3) |
| Any housekeeping/saccharolytic function (union) | 107 | 63 | 5 | — | 44.1% of all 143 control markers |

Screening the annotated Case-marker genes recovered a coherent virulence programme that is the hallmark of the dysbiotic subgingival community (**Table 3**). In total, 705 genes on 573 of 1,999 Case markers (28.7%) encoded recognised virulence/host-interaction functions across pathogenic taxa. The dominant category was proteolysis (287 genes), including a collagenase (*prtC*), thiol protease/haemagglutinin (*prtT*), dipeptidyl-peptidases (*dpp*), C-terminal processing proteases (*prc*), and S1-family peptidases, accompanied by an extensive haem/iron-acquisition repertoire (166 genes: *hmuR*, *ragA*, *tonB2*, *feoB*) and a near-complete Type IX secretion system (T9SS/PorSS; 113 genes: *porU*, *porV*, *porQ*, *porX*, *sov*), the Bacteroidota apparatus that exports gingipain-like proteases and adhesins [36, 37]. Additional functions comprised lipid A/LPS biosynthesis, gliding motility/chemotaxis, fimbrial adhesins, and multidrug efflux, with *T. forsythia*, *P. endodontalis*, *P. gingivalis,* and *T. denticola* the principal contributors. The convergence of surface proteolysis, haem acquisition and Type IX secretion — the canonical *P. gingivalis*/*T. forsythia*/*T. denticola* virulence axis — on the disease markers indicates that the discriminating features capture biologically meaningful pathogenic functions [38].

**Table 3.** Virulence/host-interaction functions encoded by periodontitis-associated (“Case”) markers (complete list in Supplementary Table 7).

| Functional category | Genes | Markers | Taxa | Representative genes | Principal contributing taxa |
| --- | --- | --- | --- | --- | --- |
| Proteolysis (proteases/peptidases) | 287 | 262 | 8 | *prtC, prtT, dpp, prc, S1B peptidase* | *T. forsythia* (196),<br>*P. endodontalis* (53),<br>*T. denticola* (14),<br>*P. gingivalis* (14) |
| Haem/iron acquisition | 166 | 157 | 5 | *hmuR, ragA, tonB2, feoB* | *T. forsythia* (150),<br>*P. endodontalis* (10),<br>*P. gingivalis* (4) |
| Type IX secretion (T9SS/PorSS) | 113 | 101 | 4 | *porU, porV, porQ, porX, sov* | *T. forsythia* (105),<br>*P. endodontalis* (6) |
| Antibiotic resistance/efflux | 62 | 60 | 9 | *norM, dinF, lolE* | *T. forsythia* (41),<br>*P. endodontalis* (13) |
| Gliding motility/chemotaxis | 49 | 35 | 8 | *gldM, gldN, sprA* | *T. forsythia* (22),<br>*T. denticola* (12) |
| LPS/lipid A biosynthesis | 46 | 41 | 5 | *lpxH, lpxK, kdsC, waaA* | *T. forsythia* (37),<br>*P. endodontalis* (5) |
| Adhesion/fimbriae | 15 | 13 | 4 | *fimA, fimbrillin-A anchor* | *P. endodontalis* (10),<br>*P. gingivalis* (3) |
| Toxin-antitoxin/stress | 10 | 10 | 4 | *hicA, abiEii-type* | *T. forsythia* (6) |
| Any virulence function (union) | 705 | 573 | 12 | — | 28.7% of 1,999 Case markers |

### 3.3 Novel candidate biomarkers beyond the established pathogens

We investigated how many periodontitis markers point to organisms or sequences not previously associated with the disease. Each marker was scored on two independent, continuous axes of novelty: (i) sequence divergence — percent identity to the closest reference genome in the Human Oral Microbiome Database (HOMD), with markers below 90% identity considered sequence-divergent; and (ii) taxonomic novelty — whether the source organism belongs to an established periodontitis-associated taxon (the Socransky red/orange complexes and widely cited modern additions such as *F. alocis*, *F. fastidiosum* and *P. endodontalis*) or to an emerging/under-recognized taxon.

Plotting each Case marker by HOMD percent identity against its log2 fold-change (periodontitis/health), with point size scaled to machine-learning feature importance, shows that the great majority of markers (1,871 of 1,999; 93.6%) fall at high sequence identity (≥90%) to a known oral genome and derive from established periodontal pathogens (**Figure 3A**, grey points), confirming that the *de novo* pipeline predominantly rediscovers canonical periodontal taxa. However, a distinct population of markers falls to the left of the 90%-identity threshold or is coloured as an emerging/under-recognized taxon: 63 sequence-divergent markers (orange) and 65 markers from emerging/under-recognized taxa (yellow), together comprising 128 markers (6.4%) that are novel on at least one axis.

**Figure 3.**
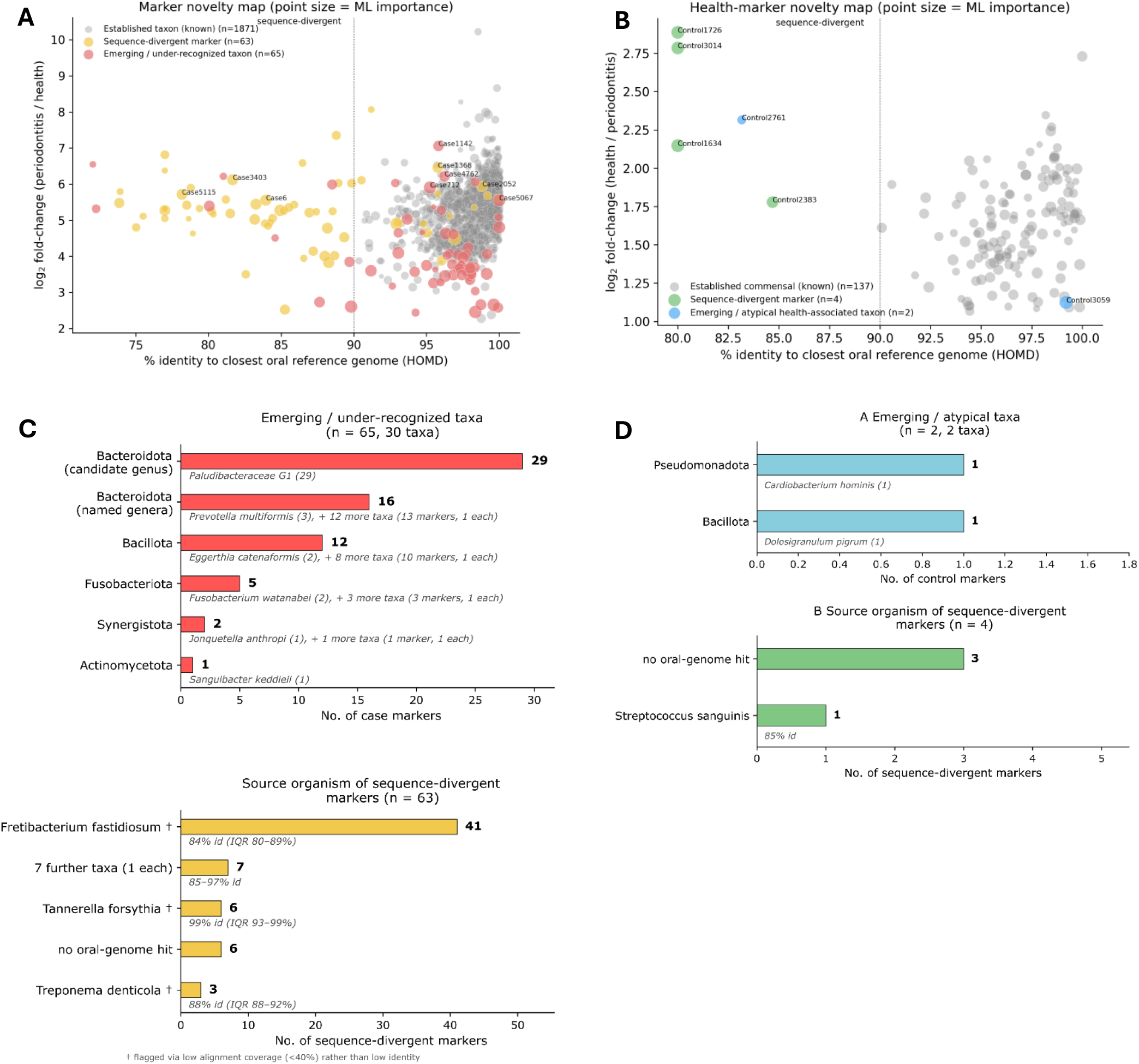
Novelty maps of the periodontitis (Case) and health (Control) markers. Percent nucleotide identity of each marker to its closest HOMD reference genome (*x*-axis) versus log₂ fold-change between groups (*y*-axis); point size indicates XGBoost feature importance (mean |SHAP|), and the dotted line marks the 90% identity threshold defining sequence-divergent markers. (A) All 1,999 Case markers: established periodontitis-associated taxon (grey, *n* = 1,871), sequence-divergent (orange, *n* = 63) or emerging/under-recognised taxon (yellow, *n* = 65). (B) All 143 Control markers: established commensal (grey, *n* = 137), sequence-divergent (blue, *n* = 4) or emerging/atypical health-associated taxon (yellow, *n* = 2). (C) Enlarged view of the 128 novel Case markers from A, labelled by source taxon (D) Enlarged view of the six novel Control markers from B, labelled by marker ID and taxon.

The largest single source of novelty was an unnamed candidate genus, *Paludibacteraceae* G1 (Bacteroidota), contributing 29 markers — a coherent, uncultivated oral taxon rarely reported in periodontitis. Other emerging taxa included the recently described species *Fusobacterium watanabei* (2 markers; named in 2020), members of the phylum Synergistetes (*Jonquetella anthropi*, *Pyramidobacter piscolens*, and highly divergent *Fretibacterium* markers), reclassified Prevotellaceae genera (*Alloprevotella rava/tannerae*, *Segatella* and *Hoylesella* spp.), and additional under-studied anaerobes [*Eggerthia catenaformis* (2 markers), *Lachnospiraceae* G8 (2), *Peptoniphilaceae* G1, *Phocaeicola abscessus*, *Anaerosphaera mikwangii* (1)] [39, 40]. The sequence-divergent markers were dominated by *F. fastidiosum*, an oral Synergistetes under-represented in reference collections, whose markers aligned to their closest oral genome at only 78–90% identity, indicating uncharacterised strains or species (**Figure 3C**).

Applying the identical screen to the 143 health (control) markers recovered fewer candidates: only 6 (4.2%) were novel, versus 128 (6.4%) among case markers, with substantially smaller effect sizes (fold-change 2.2–7.4 vs 39–163; **Figure 3B**). Two markers derived from taxa outside the established health-commensal set: *Dolosigranulum pigrum* (Control2761), an increasingly recognised member of the healthy nasal/respiratory microbiota, and *Cardiobacterium hominis* (Control3059), a HACEK-group oral commensal, at 91.2% and 99.2% identity respectively. The remaining four were sequence-divergent: three (Control1726, Control3014, Control1634) had no match to any oral reference genome and encoded exclusively mobile-genetic-element functions (IS4/IS200-family transposases, an integrase core domain) rather than taxon-defining genes, suggesting horizontally-transferred elements rather than novel commensal taxa; the fourth (Control2383, 84.7% identity) was a divergent *Streptococcus sanguinis* marker encoding the ribonucleotide reductase subunits *nrdE*/*nrdF* (**Figure 3D)**. Unlike the disease side, where novelty pointed to biologically coherent under-studied pathogens, novel health-marker candidates were sparse, weakly discriminative, and largely attributable to mobile elements. Full identities, functional annotations, and effect sizes for all novel Case and Control markers are provided in **Supplementary Table 8** and **Supplementary Table 9**, respectively.

### 3.4 Machine-learning classifier for periodontitis

We next asked whether marker abundances support accurate, transferable classification of periodontitis. Eight classifiers were trained on log-abundance features from 305 samples across seven studies and evaluated on three fully held-out external cohorts (36 samples: 17 periodontitis, 19 health) that contributed neither markers nor training data.

All eight models generalised well to the unseen test set, with ROC-AUC ranging from 0.83 (k-NN) to 0.96 (XGBoost and gradient boosting), and every model outperforming a random classifier by a wide margin (**Figure 4A, Supplementary Figure 1**). Boosted-tree ensembles performed best overall: XGBoost and gradient boosting each reached AUC = 0.96, followed closely by random forest (AUC = 0.93) and LightGBM (AUC = 0.92), with logistic regression, extra-trees (Extremely Randomized Trees), SVM-RBF [support vector machine with a Radial Basis Function (RBF) kernel] and k-NN (k-nearest neighbours algorithm) spanning AUC = 0.83–0.90. Precision–recall analysis showed consistent rankings, with average precision (AP) of 0.96 for XGBoost and 0.94 for gradient boosting, compared with a class-balance baseline of AP = 0.47, indicating that strong performance was not driven by class imbalance in the test set (**Figure 4B**).

**Figure 4.**
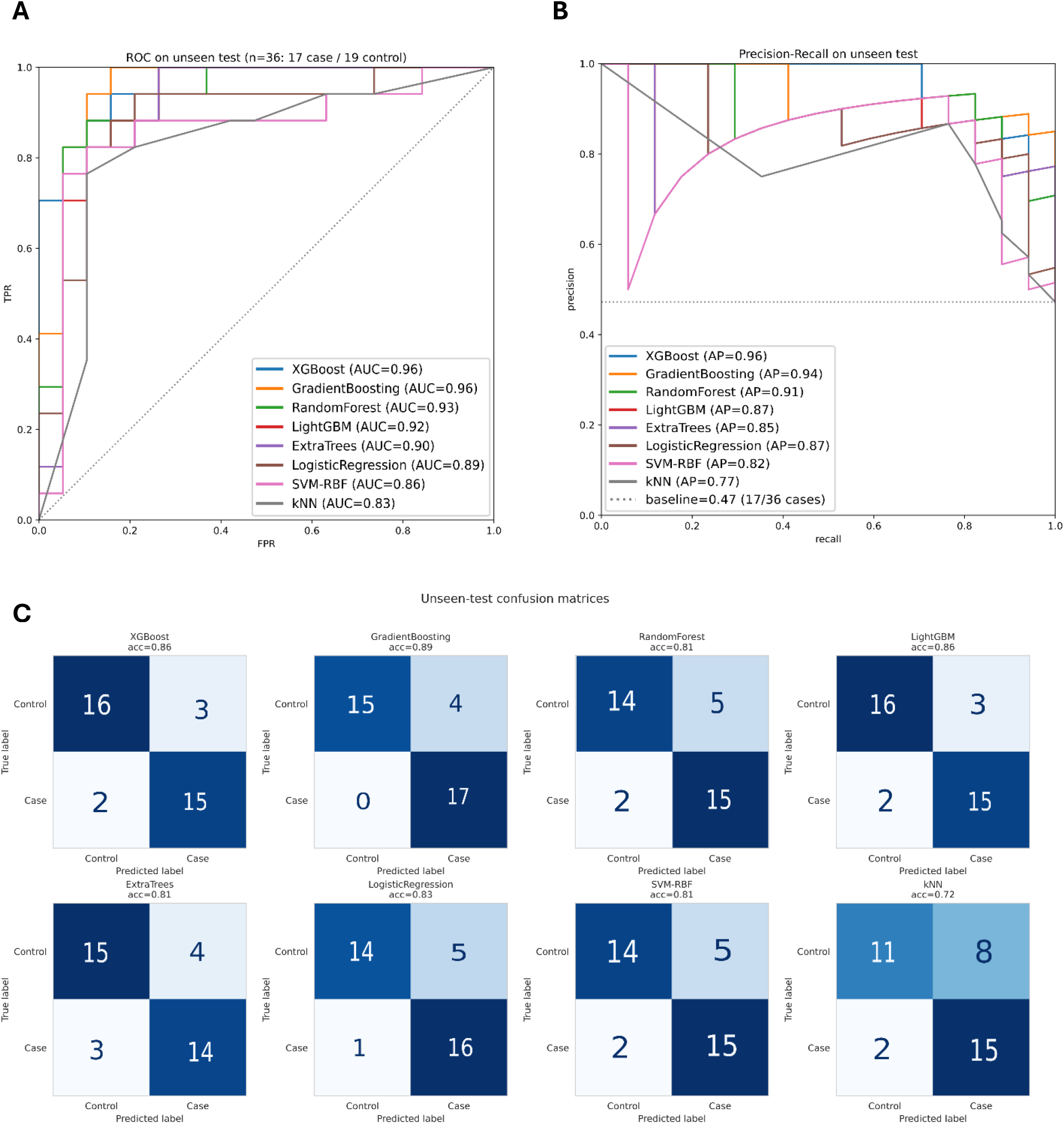
Classifier performance on the unseen external test cohorts (n = 36). (A) ROC curves for all eight models. (B) Precision–recall curves for all eight models, with class-balance baseline (AP = 0.47) shown as a dotted line. (C) Confusion matrices for all eight models on the unseen test set, annotated with overall accuracy.

Confusion matrices on the unseen test set showed balanced performance across both clinical groups for the top models (**Figure 4C**). Gradient boosting achieved the highest overall accuracy (0.89), correctly classifying all 17 periodontitis samples and miscalling only 4 of 19 controls; XGBoost and LightGBM each reached 0.86 accuracy with 2 periodontitis samples and 3 controls misclassified. Random forest, extra-trees, logistic regression, and SVM-RBF all achieved accuracy of 0.81–0.83, with errors distributed across both classes rather than concentrated in one group, while k-NN showed the weakest and most one-sided performance (accuracy 0.72, with 8 of 19 controls misclassified as periodontitis, **Supplementary Table 10**). Cross-validation within the training pool (5-fold, stratified) gave per-fold AUC in the range of approximately 0.85–0.94 across models, broadly consistent with, and in most cases slightly higher than, performance on the fully independent held-out cohorts (**Supplementary Figure 1A**). Direct comparison of training, cross-validation, and unseen-test AUC and accuracy showed the expected pattern of near-perfect training performance (AUC = 1.00 for most models), a moderate drop under cross-validation, and a further modest drop on the external test set — with boosted-tree models (XGBoost, gradient boosting, LightGBM) showing the smallest train-to-test gap, supporting their selection as the primary models for downstream interpretation (**Supplementary Figure 1B**).

### 3.5 A compact, SHAP-ranked marker panel drives model predictions

To determine whether classifier decisions rested on biologically interpretable markers rather than only statistical patterns, we computed SHAP values for the best-performing XGBoost model on the training set (n = 305) and ranked markers by their impact on model output (**Figure 5**). The top 20 markers by mean absolute SHAP value were drawn from a small number of source taxa on each side of the disease axis. Periodontitis-associated features were dominated by *Tannerella forsythia* (Case2239, Case3081, Case1245, Case402, Case1373), *Porphyromonas gingivalis* (Case3307, Case457, Case3857, Case3457, Case781, Case300), and *Treponema denticola* (Case1101, Case2679, Case3649), for which high feature values (red) consistently pushed SHAP values positive, increasing the predicted probability of periodontitis. Conversely, health-associated features were dominated by *Rothia dentocariosa* (Control289, Control806, Control1839, Control1640) and *S. mitis* (Control665, Control679), for which high feature values pushed SHAP values negative, decreasing the predicted probability of disease. For every one of these top 20 markers, the direction of the SHAP effect matched the marker’s known clinical association — red/orange-complex pathogen markers increased predicted disease risk, and commensal markers decreased it — with no instances of a marker acting in the opposite direction to its taxonomic label. This one-to-one correspondence between SHAP sign and biological identity indicates that the classifier learned the same dysbiotic signal that defines the marker set itself, rather than exploiting cohort-specific or technical confounders. Because this top-20, SHAP-ranked panel concentrates the model’s discriminative power into a small, biologically coherent and strain-conserved set of markers spanning just five source taxa (*T. forsythia*, *P. gingivalis*, *T. denticola*, *R. dentocariosa*, *S. mitis*), it constitutes a directly actionable candidate panel for a targeted, low-cost diagnostic assay, complementing the full 2,142-marker set used for genome-wide discovery and characterisation. We trained the same models using only these 20 markers and achieved robust discriminative performance (**Supplementary Figure 2**), demonstrating that this compact panel retains the essential diagnostic signal of the full marker set while offering substantial advantages for clinical translation.

**Figure 5.**
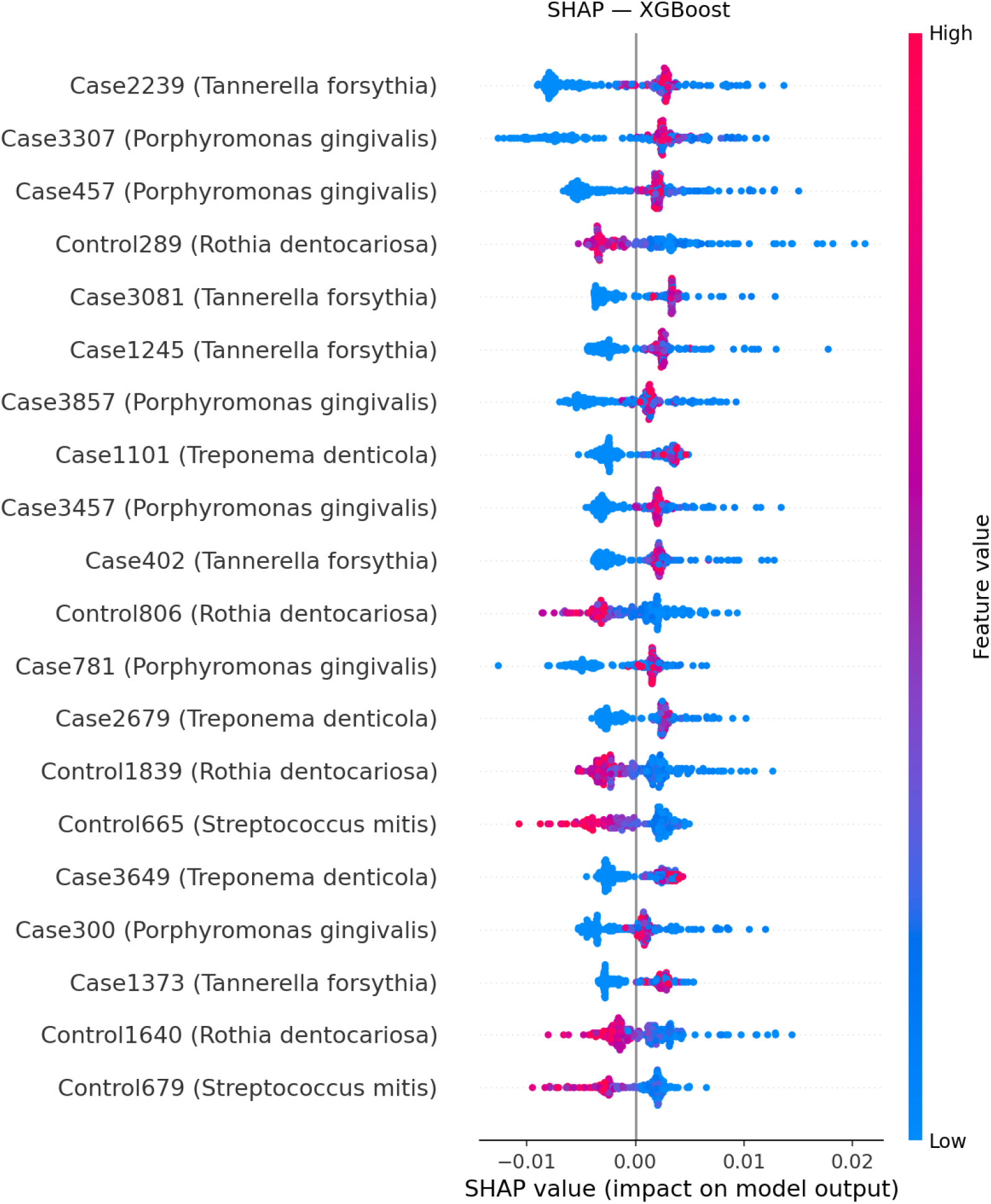
SHAP summary plot for the top 20 markers of the best-performing XGBoost model, ranked by mean absolute SHAP value. Each point represents one sample; position along the x-axis indicates the marker’s impact on model output (positive = higher predicted probability of periodontitis), and colour indicates the marker’s feature value (log-abundance) from low (blue) to high (red/pink). Markers are labelled by marker ID and source taxon.

### 3.6 Strain-conservation confirms markers are genuinely conserved genomic features

To confirm that markers correspond to genuine, generalisable genomic features rather than strain-specific or assembly artifacts, the twenty most important markers were aligned by BLAST against the NCBI nucleotide database and their conservation examined across strains of their assigned species (**Supplementary Figure 3**). Thirteen markers had at least one subject strain covering >90% of their length, and over the aligned region, all were highly conserved (≈92–99% mean nucleotide identity) across multiple strains of the target species. For example, the *T. forsythia* marker Case2239 aligned essentially end-to-end (query coverage 100%) at ∼99.4% mean identity across three *T. forsythia* strains; comparable strain-level conservation was observed for markers of *P. gingivalis* (Case3307; 31 strains >90% coverage), *T. denticola* (Case1101), and the health-associated *Rothia dentocariosa* (Control289). This strain-level conservation explains why the markers are detectable across independent cohorts regardless of which strain of a given species is present.

## 4. Discussion

This study set out to determine whether a reference-free, *de novo* marker-discovery strategy could both recover established periodontal microbiology and uncover biomarkers invisible to conventional, database-dependent pipelines, and whether such markers could support an accurate, externally validated, non-invasive classifier. The results address all four aims. First, the 2,142 markers recovered by MetaMarker, identified without any a priori mapping to a reference database, cleanly reconstructed the canonical red/orange-complex dysbiotic signature—elevation of *T. denticola*, *T. forsythia* and *P. gingivalis* in disease and of *S. mitis*, *S. oralis* and *R. dentocariosa* in health—at both the individual-marker and aggregated-organism level. This convergence with decades of culture-, 16S-, and shotgun-based periodontal microbiology is an important internal validation of the approach.

Second, and most novel, 128 periodontitis markers (6.4%) pointed to organisms that are either sequence-divergent from the Human Oral Microbiome Database or belong to taxa rarely or never implicated in periodontitis, most prominently an uncultivated Paludibacteraceae genus and divergent *F. fastidiosum* strains. Third, the functional annotation of Case markers revealed a coherent virulence programme—proteolysis, haem/iron acquisition, and a near-complete Type IX secretion system—concentrated in *T. forsythia*, *P. endodontalis*, *P. gingivalis* and *T. denticola*. That nearly 29% of Case markers carried a recognisable virulence or host-interaction function, despite markers being selected purely on statistical differential abundance with no functional criterion, indicates that the discriminative signal captured in our biomarkers is not an arbitrary sequence signature but tracks genuine pathogenic machinery. The T9SS/PorSS component is particularly notable because this secretion apparatus is specific to the Bacteroidota and mechanistically exports the gingipain-family proteases and adhesins long implicated in periodontal tissue destruction; its recovery here, entirely from unsupervised marker discovery, provides an independent line of evidence for its centrality to the disease process.

Fourth, the machine-learning results show that marker abundance alone—without clinical, host, or demographic covariates—is sufficient for accurate, generalisable classification. The strong performance of boosted-tree models (AUC = 0.96) on three fully independent, geographically and technically distinct external cohorts, none of which contributed to marker discovery or training, is a meaningfully stronger test of generalisability than the within-cohort cross-validation reported by many prior microbial-biomarker studies. The modest, consistent train-to-cross-validation-to-external-test performance drop, smallest for the boosted-tree models, further supports these models as the most robust for translational use. Critically, the SHAP analysis showed that the classifier’s top 20 features were drawn from the same taxa that drove the unsupervised differential abundance analysis, with each marker’s SHAP direction matching its known clinical association.

Taken together, these results argue for two complementary translational directions. The full 2,142-marker set is a discovery resource, offering candidate taxa and genes for follow-up culturing, metatranscriptomic, or functional validation, particularly for the uncultivated Paludibacteraceae and divergent Fretibacterium lineages. The compact, SHAP-ranked, 20-marker panel spanning only five taxa is a candidate diagnostic resource, small enough to be plausibly implemented as a targeted PCR- or probe-based assay on plaque, avoiding the cost and turnaround time of shotgun sequencing while retaining the discriminative power of the full marker set.

A limitation of this study is its cross-sectional, case–control design, so markers—however robust—reflect association with periodontitis status at sampling, not causation or temporal sequence in disease progression; longitudinal cohorts would be needed to determine whether marker abundance changes precede, accompany, or follow clinical attachment loss. A further limitation is that potential confounders known to influence the oral microbiome—smoking status, diabetes, antibiotic use, and periodontitis stage/grade—were not available for harmonised analysis across all ten BioProjects and could not be adjusted for. Future work should prioritize culturing or targeted long-read sequencing of the uncultivated Paludibacteraceae genus and divergent Fretibacterium lineages to resolve their genomic content and confirm pathogenic potential; longitudinal sampling to establish whether novel markers change ahead of clinical disease onset; incorporation of host-response biomarkers alongside the microbial panel to test complementarity; and prospective clinical validation of the 20-marker panel in a point-of-care format across more diverse populations and periodontitis severities than represented here.

## Data availability

All the data used in this study are publicly available and mentioned in Table 1. The MetaMarker tool we used to discover the markers is also available at https://github.com/ClinicalAI/MetaMarker/.

## Funding sources

There is no funding for this study.

## Conflict of Interest Statement

All authors declare no conflicts of interest.

## Ethics Statement

This study is a meta-analysis based on published data. All data analysed were obtained from publicly available literature, and no original animal or human experiments were conducted. Therefore, ethical approval and informed consent were not required.

## Credit authorship contribution statement

**Mohamad Koohi-Moghadam:** Conceptualization, Software, Methodology, Investigation, Data curation, Writing – review and editing, Writing – original draft, Resources, Project administration, Formal analysis, Visualization, Validation, Supervision. **Wai Keung Leung:** Writing – review and editing, Investigation, Data curation, Formal analysis.

## Supplementary materials

Supplementary material associated with this article can be found in the attached file.

## Notes

### Competing Interest Statement

The authors have declared no competing interest.

